# Real-Time Kinematic Adaptive Deep Brain Stimulation Safely Reduces Gait Impairment and Freezing of Gait in Parkinson’s Disease

**DOI:** 10.64898/2026.02.23.26346487

**Authors:** Shreesh Karjagi, Yasmine M. Kehnemouyi, Matthew N. Petrucci, Laura Parisi, Emilia F. Lambert, Jillian A. Melbourne, Pranav Akella, Kevin B. Wilkins, Johanna O’Day, Hannah J. Dorris, Cameron Diep, Aryaman S. Gala, Chuyi Cui, Shannon L. Hoffman, Prerana Acharyya, Jeffrey A. Herron, Helen M. Bronte-Stewart

## Abstract

Gait impairment (GI) and freezing of gait (FOG) affect 80% of patients with advanced Parkinson’s disease. Continuous deep brain stimulation (cDBS) provides limited adaptability to address the episodic nature of FOG due to fixed parameters. Neural biomarkers for adaptive DBS are limited by signal artifacts and poor FOG classification. Wearable inertial measurement units (IMUs) offer a promising alternative by directly measuring signatures of GI&FOG. We developed Kinematic adaptive DBS (KaDBS), the first intelligent system to dynamically modulate stimulation in response to real-time gait metrics. KaDBS integrates bilateral shank-mounted IMUs with an investigational neurostimulator through a wireless architecture enabling step-detection, arrhythmicity calculation, and probabilistic FOG classification. Two control algorithms were implemented: an arrhythmicity model based on stride variability, and a P(FOG) classifier implementing tri-state control based on stepwise freezing probabilities. In the largest KaDBS cohort to date (n=8), we compared OFF, cDBS, KaDBS, and intermittent DBS during harnessed stepping and free walking. KaDBS was safe and well tolerated with no serious adverse events; symptom-free reports were 87.5% and 71.4% for arrhythmicity and P(FOG) models respectively, compared to 50.0% for cDBS. All symptoms were mild, transient, and resolved without intervention. KaDBS significantly reduced percent time freezing versus OFF during stepping-in-place (35.8%, P= 4.80 × 10⁻³) and free walking (33.4%, p = 9.00 × 10⁻⁴). Therapeutic effects concentrated in baseline freezers: two participants with 100% time freezing during OFF achieved complete resolution with KaDBS, while non-freezers maintained stable gait. These findings establish KaDBS as a safe, effective approach to personalized neuromodulation for PD.

## Introduction

Gait impairment in Parkinson’s disease (PD) manifests as a complex phenomenon characterized by slow movements, arrhythmic stepping, and episodic freezing of gait (GI&FOG) - affecting up to 80% of advanced-stage participants (*1*). Multiple neural networks contribute to these complex symptoms(*1–3*), and while continuous deep brain stimulation (cDBS) provides therapeutic benefits for cardinal motor symptoms(*4–6*), and improves GI&FOG to a certain degree in individuals with PD who are responsive to medication(*5*), its impact remains limited. Over time, non-dopaminergic contributions to GI&FOG emerge as disabling symptoms refractory to both dopaminergic medication and DBS in dopaminergic networks(*7*), necessitating more precise biological interactions with these complex neural networks. This symptomatic evolution is further constrained by the current paradigm: fixed stimulation parameters are not designed to adapt to the episodic nature of FOG and the progression of FOG severity(*8, 9*), and programming protocols traditionally prioritize improvement of tremor, bradykinesia, and rigidity with less consistent emphasis on gait and axial symptoms(*10, 11*). These limitations have motivated the development of adaptive deep brain stimulation (aDBS), which modulates stimulation parameters in real time in response to pathophysiological biomarkers for GI&FOG in PD.

While neural biomarkers derived from local field potentials (LFPs), including subthalamic nucleus (STN) beta-band power(*12–15*), beta-band burst durations(*16, 17*),and cortico-basal finely tuned gamma(*18, 19*), have demonstrated utility as control signals for aDBS, their implementation poses particular challenges for monitoring dynamic gait states. These approaches require specialized sense-friendly stimulation configurations, are susceptible to stimulation and movement related artifacts, and provide lower fidelity signals compared to direct behavioral measurements for classifying complex gait behaviors such as FOG episodes, where rapid and accurate detection is crucial for therapeutic response(*20*). Furthermore, the spatially distributed nature of STN activity patterns during FOG states creates additional challenges for single-site recording approaches(*21, 22*). The limitations of neural biomarkers have prompted exploration of behavioral inputs recorded from peripheral sensors as alternative control signals for kinematic (K)aDBS for GI&FOG.

Wearable inertial measurement units (IMUs) offer a practical approach for detecting gait abnormalities with high temporal resolution, capturing the characteristic kinematic signatures of GI&FOG(*23–25*). These sensors can be deployed in ambulatory settings, allowing symptom monitoring during activities of daily living where GI&FOG most commonly manifest(*26–28*). Furthermore, the growing commercial availability and decreasing cost of miniaturized wearable technology makes sensor-based approaches increasingly practical for long-term therapeutic applications, potentially enabling more accessible personalized neuromodulation systems that directly target GI&FOG(*29, 30*). This approach has shown promise in early demonstrations, first for resting tremor(*31*), then for gait in benchtop testing(*32*), and recently showed efficacy of KaDBS for GI&FOG in a single-participant case study using an arrhythmicity-based control algorithm(*33*).

Here we present the first clinical trial of KaDBS that safely modulated neurostimulation in response to behavioral inputs relevant to GI&FOG in PD. In the largest KaDBS cohort to date (n=8), we conducted a randomized double-blind study evaluating the safety, tolerability, and efficacy of two novel control strategies: one based on a single gait arrhythmicity threshold and another using probabilistic FOG detection. We developed an intelligent distributed architecture integrating kinematic data streams from shank-mounted IMUs with an implanted neurostimulator via a PC in the loop to automatically adjust DBS during both harnessed stepping and naturalistic free walking tasks. This first-of-its-kind investigation demonstrated robust adaptive neuromodulation, which adjusted neurostimulation amplitude automatically in response to relevant kinematic signals of impaired gait. The results of this study provide a significant step toward personalized therapy for such debilitating symptoms of Parkinson’s disease.

## Results

### Demographic Characteristics

Eight participants with Parkinson’s disease were enrolled in this study (6 men, 2 women; mean age 68.4 ± 5.2 years). All participants exhibited moderate to severe motor symptoms (MDS-UPDRS III: 37.8 ± 7.5 in the preoperative off-medication state) and had long disease durations (15.4 ± 2.4 years). Seven participants (87.5%) reported freezing of gait (FOG-Q score > 1), and one participant was a non-freezer with mild gait impairment (MDS-UPDRS III Gait =1. All participants received the Summit RC+S deep brain stimulation system (Medtronic, Inc): for five participants it was their first implanted neurostimulator and for three it was a replacement neurostimulator (table 1).

**Table 1:**
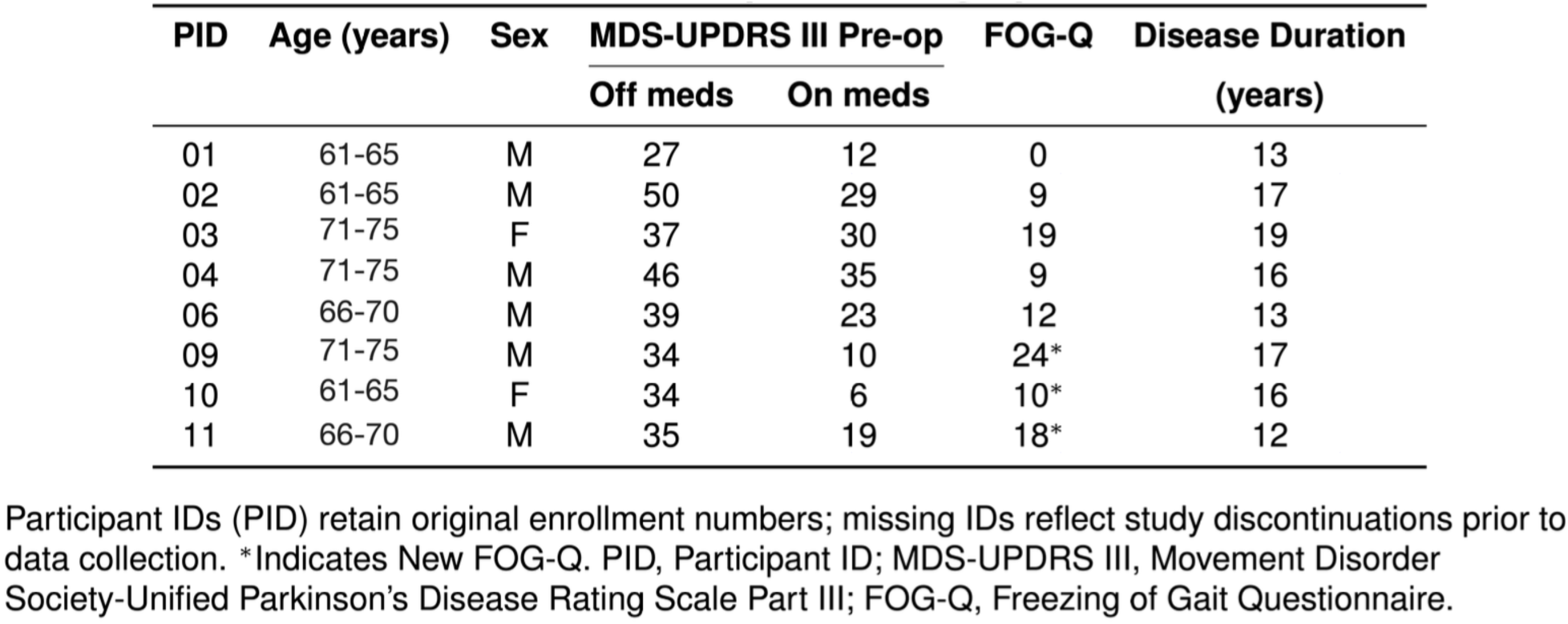
Participant Demographics

Participants underwent two testing visits in a bench-to-bedside progression separated by three months (fig. S1). Visit 1 established KaDBS safety during a harnessed stepping-in-place (SIP) task performed on force platforms, where participants were safely supported throughout testing. Following successful safety demonstration, visit 2 progressed to naturalistic free walking in a turning and barrier course (TBC) with dividers simulating real-world environments including narrow hallways and doorways. All assessments occurred in the practically defined off-medication state, and four randomized, double-blind stimulation conditions were tested at each visit: no stimulation (OFF), conventional continuous DBS (cDBS), kinematic adaptive DBS (KaDBS), and random intermittent DBS (iDBS).

### Distributed Kinematic Adaptive DBS System

The KaDBS system integrated bilateral shank-mounted IMUs with an investigative sensing-enabled neurostimulator (Summit RC+S, Medtronic, Inc) through a distributed wireless architecture (Fig. 1A). The system established two wireless communication pathways: a Bluetooth connection (12m range) between the shank IMUs and the PC-based processing unit, and RF telemetry (1m range) between the clinician telemetry module and the implanted neurostimulator.

**Fig. 1.**
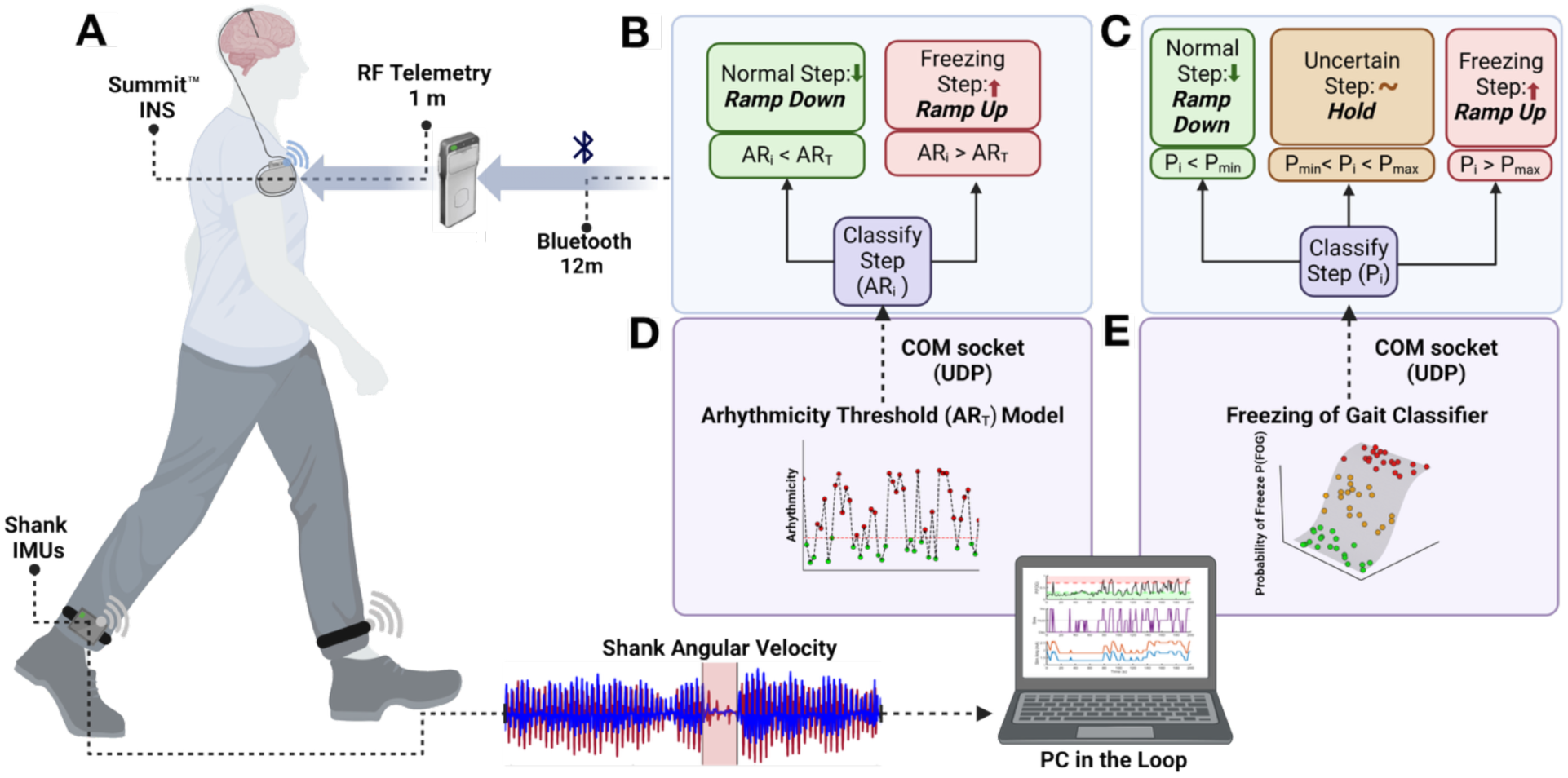
Schematic of the distributed KaDBS system. (**A**) The distributed system integrated bilateral shank-mounted inertial measurement units (IMUs) with a sensing-enabled neurostimulator (Summit RC+S) through wireless telemetry. (**B, D**), The arrhythmicity threshold (ARt) model modulated stimulation amplitude based on detected stride variability: ramping down when arrhythmicity (ARi) was below threshold (ARt) and ramping up when ARi exceeded threshold. (**C, E**), The freezing of gait classifier estimated stepwise freezing probabilities (Pi), where stimulation amplitude decreased during normal stepping (Pi < Pmin), stayed at the same amplitude when uncertain (Pmin ≤ Pi ≤ Pmax), and increased during FOG (Pi > Pmax ). The PC-based Gait Processing Application analyzed bilateral shank angular velocity signals in real-time and communicated stimulation state changes through a COM socket (UDP) interface (Summit RC+S neurostimulator and communicator images credited to Medtronic, Inc).

The IMUs captured shank angular velocity during gait, generating characteristic oscillatory signals that reflect stepping movements. These kinematic signals streamed to a PC-based Gait Processing Application that performed real-time step detection, gait parameter computation, and stimulation control, then communicated adjustments to the neurostimulator through a UDP interface.

We implemented two novel control algorithms for stimulation modulation (Fig. 1, B to E). The arrhythmicity threshold (AR_t_) model provided binary control based on stride variability, ramping stimulation amplitude down when the measured arrhythmicity (AR_i_) fell below the participant-specific threshold (AR_i_ < AR_t_), and ramping amplitude up when AR_i_ exceeded the participant-specific threshold (AR_i_ > AR_t_) (Fig. 1, B and D). Participant specific arrhythmicity thresholds were determined during initial calibration based on individual gait characteristics. The freezing of gait classifier implemented a tri-state dual threshold control algorithm based on computed step-wise freezing probabilities (P_i_): ramping stimulation amplitude down during normal stepping (P_i_ < P_min_), maintaining current stimulation amplitude when classification was uncertain (P_min_ ≤ P_i_ ≤ P_max_), and ramping stimulation amplitude up when freezing was likely (P_i_ > P_max_) (Fig. 1, C and E). Typical probability thresholds were set at P_min_ = 0.3 and P_max_ = 0.7. The control algorithms implemented asymmetric ramp rates: amplitude increases occurred faster than decreases to prevent rapid stimulation withdrawal following FOG resolution, which could precipitate symptom recurrence.

### Quantitative Kinematic Adaptive DBS Parameter Calibration

KaDBS requires a participant-specific therapeutic stimulation window that defines the permissible range for amplitude adaptation, bounded by the minimum amplitude providing clinical benefit (Imin) and the maximum amplitude tolerated without side effects (Imax). To establish this window, we evaluated each participant’s gait performance across multiple stimulation amplitudes normalized to the stimulation amplitude used clinically (100%). During the SIP visit, we analyzed bilateral force plate data, while for the TBC visit, we analyzed shank angular velocity from IMU recordings. An example in one participant during the SIP task is shown in Fig. 2, A and B. Force traces were normalized to body weight, where 100% indicated full weight-bearing and 0% indicated foot-off. At 0% stimulation amplitude, the participant exhibited severe gait impairment with 75.2% time freezing during the 30-second trial (Fig. 2A). Freezing decreased to 4.2% at 50% clinical amplitude and 2.5% at 75% clinical amplitude, with complete resolution between 85-115% clinical amplitude (Fig. 2B). This systematic assessment of freezing behavior across stimulation amplitudes led to defining the therapeutic window between Imin (75%) and Imax (100%) clinical stimulation amplitude, establishing the allowable range for stimulation amplitude to adapt during KaDBS.

**Fig. 2.**
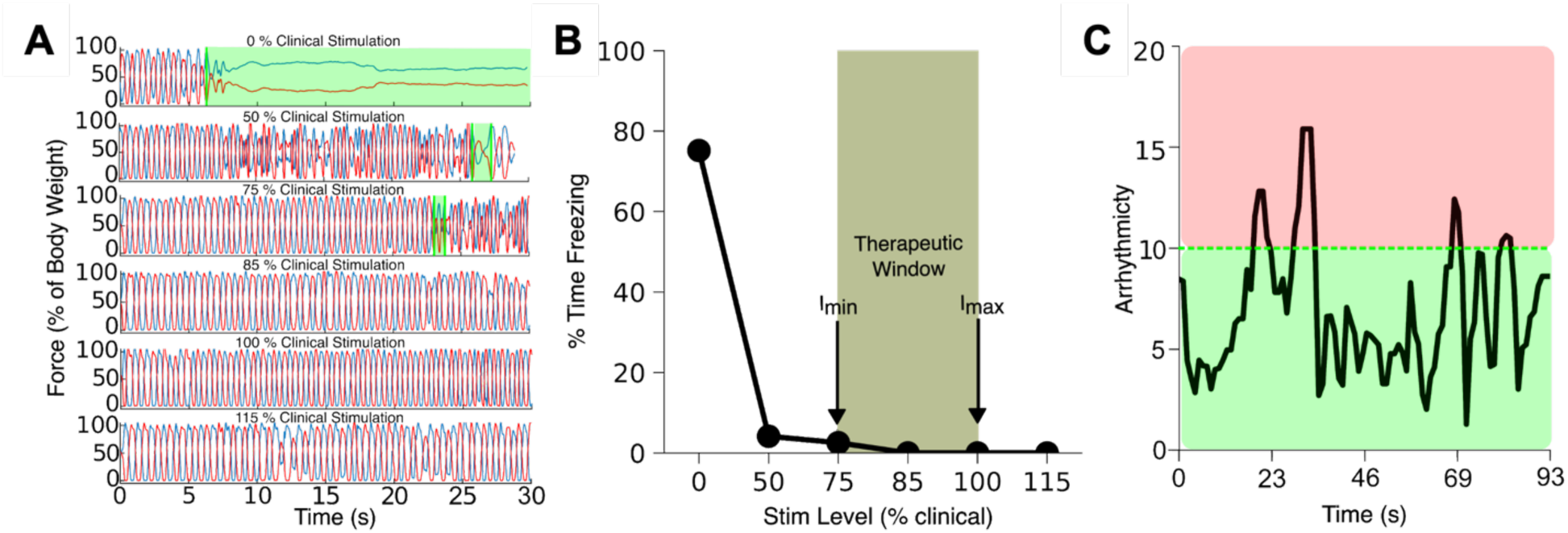
Quantitative KaDBS parameter calibration. (**A**) Performance during SIP task at varied stimulation amplitudes (0-115% of clinical amplitude). Vertical ground reaction forces shown as percentage of body weight. (**B**) Percentage time spent freezing plotted against stimulation levels, with therapeutic window (olive green) indicating allowable stimulation range. (**C**) Arrhythmicity time series during SIP task establishing participant-specific threshold ARt (dotted line) at 85% of average arrhythmicity, distinguishing high (red) and low (green) arrhythmicity periods for KaDBS implementation. Data in (**A**) and (**B**) from one participant, (**C**) from a different participant.

To determine the participant-specific AR_t_ for KaDBS, we analyzed gait metrics during the same stimulation titrations used to establish the therapeutic stimulation amplitude window. Fig. 2C shows the gait arrhythmicity time series from another representative participant during the SIP task across various stimulation amplitudes. We set AR_t_ at 85% of the mean arrhythmicity observed during these trials, as this value optimally distinguished between normal stepping (arrhythmicity below threshold, green) and arrhythmic gait patterns that typically preceded freezing episodes (arrhythmicity above threshold, red).

Individual KaDBS parameters derived from these calibration procedures are summarized in table 2, showing participant-specific therapeutic windows and thresholds for both algorithms across the two testing visits.

**Table 2:** KaDBS parameters

| PID | Visit | STN Contacts |  | STN Range (mA) |  | Ramp Up/Down (mA s<sup>-1</sup>) |  | Arrhythmicity Threshold |
| --- | --- | --- | --- | --- | --- | --- | --- | --- |
|  |  | LSTN | RSTN | LSTN | RSTN | LSTN | RSTN |  |
| 1 | SIP | 1<sup>-</sup>, 2<sup>-</sup> | 9<sup>-</sup> | 3.4–4.6 | 2.0–2.7 | 0.125/0.0625 | 0.125/0.0625 | 6.5 |
| 1 | TBC | 1<sup>-</sup>, 2<sup>-</sup> | 9<sup>-</sup> | 3.5–4.6 | 2.0–2.7 | 0.125/0.0625 | 0.125/0.0625 | † |
| 2 | SIP | 0<sup>-</sup>, 1<sup>-</sup> | 8<sup>-</sup>, 9<sup>-</sup> | 3.6–4.7 | 6.2–6.2 | 0.0625/0.0625 | 0/0 | 9.0 |
| 2 | TBC | 0<sup>-</sup>, 1<sup>-</sup> | 8<sup>-</sup>, 9<sup>-</sup> | 3.6–4.7 | 6.2–6.2 | 0.0625/0.0625 | 0/0 | † |
| 3 | SIP | 1<sup>-</sup>, 2<sup>-</sup> | 9<sup>-</sup> | 2.0–2.6 | 2.2–2.9 | 2.4/0.125 | 2.8/0.125 | 10.0 |
| 3 | TBC | 1<sup>-</sup>, 2<sup>-</sup> | 9<sup>-</sup> | 2.2–2.9 | 2.3–3.1 | 2.8/0.125 | 3.2/0.125 | † |
| 4 | SIP | 1<sup>-</sup> | 10<sup>-</sup> | 1.4–3.4 | 1.8–4.2 | 0.25/0.125 | 0.25/0.125 | 10.0 |
| 4 | TBC | 1<sup>-</sup> | 10<sup>-</sup> | 2.2–2.9 | 2.8–3.7 | 2.8/0.125 | 3.6/0.125 | † |
| 6 | SIP | 1<sup>-</sup>, 2<sup>-</sup> | 8<sup>-</sup>, 9<sup>-</sup> | 5.4 | 5.2–5.8 | 0/0 | 0.0375/0.01875 | 25.0 |
| 9 | SIP | 2<sup>-</sup> | 9<sup>-</sup> | 1.8–2.4 | 1.3–1.7 | 2.4/0.125 | 1.6/0.125 | 10.0 |
| 9 | TBC | 2<sup>-</sup> | 9<sup>-</sup> | 1.2–2.6 | 0.9–1.9 | 0.25/0.125 | 0.25/0.125 | † |
| 10 | SIP | 2<sup>-</sup> | 8<sup>-</sup>, 9<sup>-</sup> | 2.6–3.4 | 4.7–6.2 | 0.2/0.1 | 0.2/0.1 | 10.0 |
| 10 | TBC | 2<sup>-</sup> | 8<sup>-</sup>, 9<sup>-</sup> | 2.6–3.4 | 4.7–6.2 | 0.2/0.1 | 0.2/0.1 | † |
| 11 | SIP | 1<sup>-</sup> | 9<sup>-</sup> | 3.0–4.0 | 3.0–4.0 | 4/0.125 | 4/0.125 | 13.0 |
| 11 | TBC | 1<sup>-</sup> | 9<sup>-</sup> | 3.1–4.1 | 3.1–4.1 | 4/0.125 | 4/0.125 | † |

All configurations used case as anode. Superscript <sup>-</sup> indicates cathode. Double dash (–) indicates amplitude range; forward slash (/) separates ramp up/down values. †TBC trials used probabilistic P(FOG) classifier (P<sub>min</sub> = 0.3, P<sub>max</sub> = 0.7) instead of arrhythmicity threshold.

### Kinematic Adaptive DBS controllers adjusted stimulation in real-time based on gait biomarkers

Following calibration and therapeutic window establishment, we implemented two control algorithms in real-time, each demonstrating unique approaches to dynamic stimulation adaptation. Fig. 3. illustrates representative data from both controllers.

**Fig. 3.**
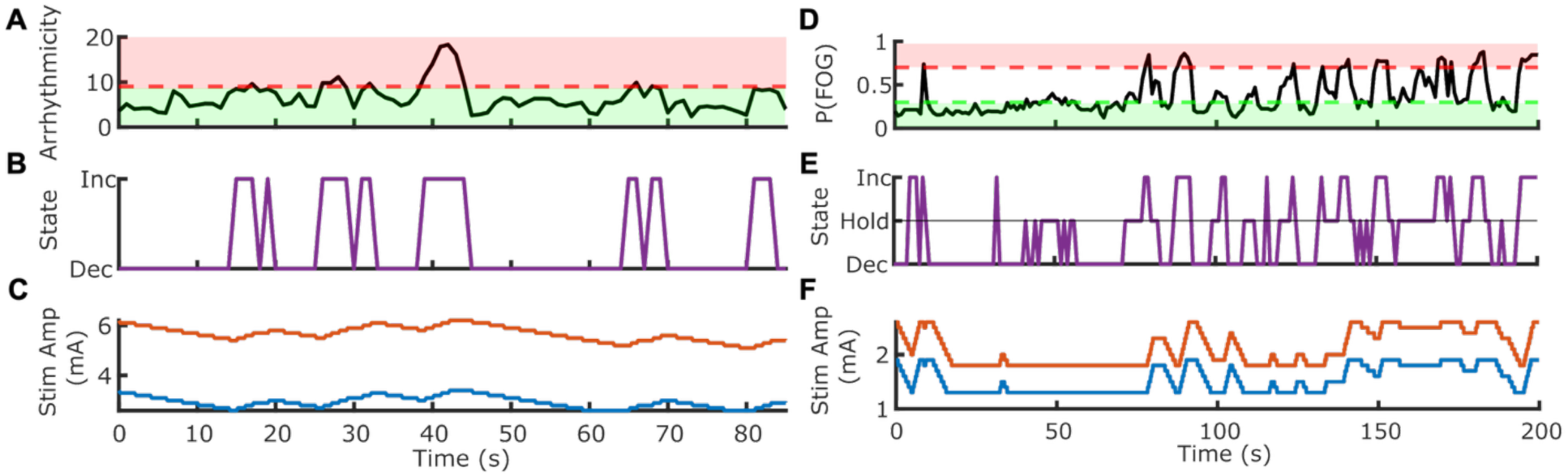
Real-time demonstration of two KaDBS control algorithms. A-C, Arrhythmicity-based control (n=1): real-time arrhythmicity measurements (**A**) with participant-specific single threshold (red dashed line), corresponding state transitions (**B**), and bilateral stimulation amplitude modulation (C) with left (blue trace) and right (orange trace) STN stimulation in mA during 80-second SIP trial. D-F, P(FOG)-based control (n=1): probability of freezing of gait (**D**) with dual thresholds (red dashed line, Pmax =0.7; green dashed line, Pmin =0.3), tri-state control implementation (**E**), and resulting stimulation amplitude adjustments (**F**) with left (blue trace) and right (orange trace) STN stimulation during 200-second free walking TBC trial. Data from two different participants, each testing one control algorithm.

The arrhythmicity-based algorithm (Fig. 3, A to C) demonstrated responsive DBS adaptation to changes in gait rhythmicity during the SIP task. Real-time arrhythmicity measurements (Fig. 3A) were continuously compared against the participant-specific AR_t_ (red dashed line). When arrhythmicity exceeded threshold at 39 seconds, the control state (Fig. 3B) transitioned to “increase,” triggering bilateral stimulation increases (Fig. 3C): RSTN (orange) from 5.8 to 6.2 mA and LSTN (blue) from 3.0 to 3.6 mA. At 63 seconds, when arrhythmicity fell below threshold, stimulation decreased to Imin (LSTN: 2.6 mA, RSTN: 5.2 mA) (movie S1).

The P(FOG)-based controller demonstrated adaptation during the free walking TBC task (Fig. 3, D to F). This algorithm continuously calculated the P(FOG) for each step, which fluctuated between 0.2-0.8 over the 200-second trial period (Fig. 3D). Control decisions were governed by dual thresholds: Pmax (0.7, upper red dashed line) and Pmin (0.3, lower green dashed line). The tri-state control system (Fig. 3E) implemented “Increase,” “Hold,” or “Decrease” states based on these threshold crossings. This produced dynamic bilateral stimulation control (Fig. 3F), with left (blue trace) and right (orange trace) STN stimulation independently modulated between 1.5-2.0 mA and 2.0-2.5 mA, respectively, in response to changing gait states (movie S2).

### Kinematic Adaptive DBS was safe and tolerable

Having demonstrated the real-time performance of both control algorithms, we next assessed their safety and tolerability across all participants. No testing sessions required termination due to adverse events. For the arrhythmicity model, 87.5% (7/8) of symptom reports indicated no adverse sensations, with one report of mild, transient imbalance (12.5%) (Fig. 4A). The P(FOG) model showed similar tolerability: 71.4% (5/7) of reports indicated no symptoms, with imbalance and dizziness each accounting for 14.3% (one report each) (Fig. 4B). By comparison, clinical cDBS resulted in only 50.0% (5/10) symptom-free reports, with dizziness (20.0%, 2/10), imbalance (10.0%, 1/10), pulling (10.0%, 1/10), and nausea (10.0%, 1/10) comprising the remainder (Fig. 4C). The higher number of cDBS reports (10) relative to participants (8) reflects multiple symptoms reported by some individuals. All symptoms were mild, transient, and resolved without intervention.

**Fig. 4.**
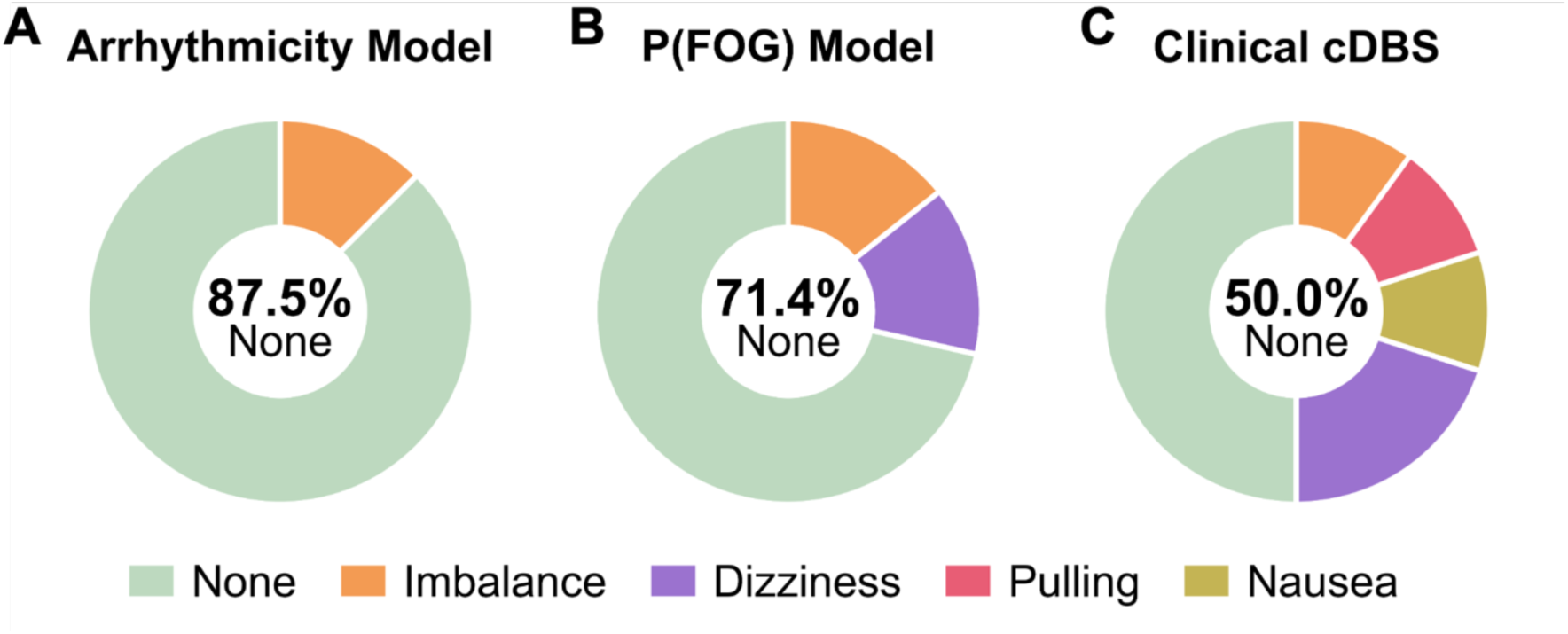
Safety assessment of KaDBS control algorithms and clinical cDBS. Participant-reported symptoms assessed after gait tasks with (**A**) arrhythmicity model (n = 8), (**B**) P(FOG) model (n = 7), and (**C**) clinical cDBS (n = 8). Percentages represent the proportion of total symptom reports; some cDBS participants reported multiple symptoms (10 total reports from 8 participants).

### Kinematic Adaptive DBS Improved Stepping in Harnessed Gait Task

In our cohort of participants with GI&FOG (n=8), we examined the effects of different DBS conditions on SIP task performance. DBS condition demonstrated a significant effect on percent time freezing (F(3,21)=5.985, p = 4.10 × 10⁻³, ηp²=0.46, n=8; Fig. 5A, table S1). Participants spent 51.35 ± 12.12% (estimated marginal mean ± SE) of time freezing during OFF DBS. All active DBS conditions produced significant reductions in percent time freezing compared to OFF: KaDBS reduced freezing by 35.83 ± 9.90% (p = 4.80 × 10⁻³), cDBS by 33.83 ± 9.90% (p = 7.80 × 10⁻³), and iDBS by 32.87 ± 9.90% (p = 9.80 × 10⁻³; table S1).

**Fig. 5.**
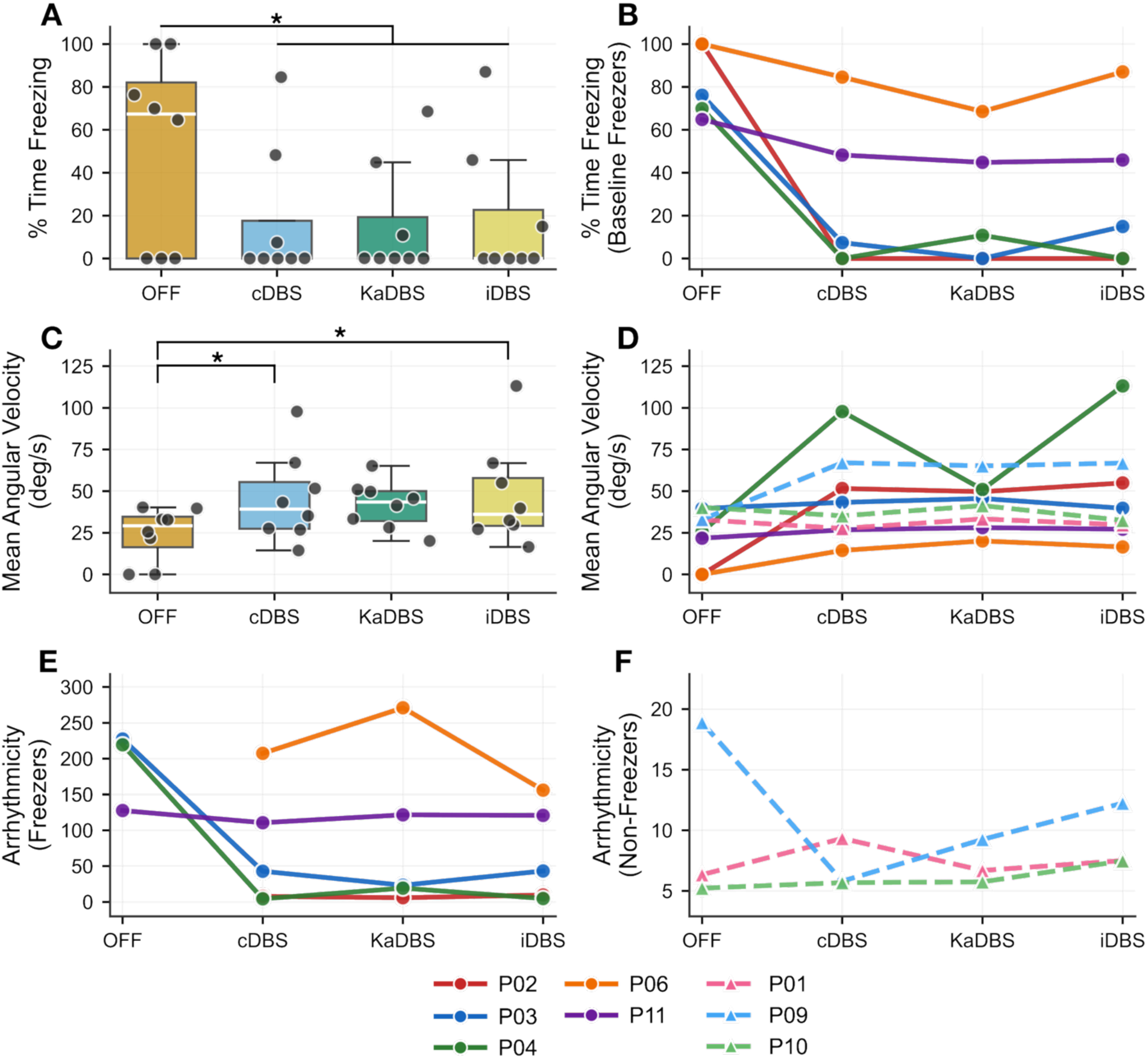
Gait metrics across stimulation conditions during SIP task. (**A**) Population % time Freezing (N=8) shown as colored boxplots with individual data points along with mean and standard deviation. * Indicates p < 0.05. (**B**) % Time freezing trends for baseline freezers. (**C**) Population mean shank angular velocity (N=8) shown as colored boxplots with individual data points along with mean and standard deviation. * Indicates p < 0.05. (**D**) Mean shank angular velocity trends for baseline freezers and for non-freezers. Arrhythmicity trends for (**E**) baseline freezers and (**F**) for non-freezers. Baseline freezers (solid lines, circles, n=5) defined as participants with >0% freezing during OFF condition, non-freezers (dashed lines, triangles, n=3) defined as participants with 0% baseline freezing.

Participants were stratified based on baseline freezing behavior into baseline freezers (>0% freezing during OFF DBS, n=5) and non-freezers (0% freezing during OFF DBS, n=3). Individual trends revealed heterogeneous treatment effects among baseline freezers (Fig. 5B). Participant 02 achieved complete elimination of percent time freezing across all DBS conditions. Participant 03 showed complete elimination of freezing selectively with KaDBS, while experiencing residual freezing with cDBS and iDBS. Participant 04 demonstrated no freezing with both cDBS and iDBS while experiencing freezing episodes during KaDBS. Participants 06 and 11 exhibited modest therapeutic benefit from DBS compared to OFF, with optimal responses observed during KaDBS. Non-freezers (Participants 01, 09, 10) maintained 0% freezing across all DBS conditions (table S2).

Mean shank angular velocity (SAV) showed a significant effect of DBS condition during SIP (F(3,21)=3.504, p = 3.33 × 10⁻², ηp²=0.33, n=8; Fig. 5C, table S1). Participants had a mean SAV of 24.129 ± 8.173 deg/s (estimated marginal mean ± SE) during OFF DBS. All DBS conditions improved mean SAV compared to OFF: cDBS by 21.3 ± 8.1 deg/s, KaDBS by 17.7 ± 8.1 deg/s, and iDBS by 23.4 ± 8.1 deg/s. Post-hoc comparisons revealed that both iDBS (p = 2.51 × 10⁻²) and cDBS (p = 4.48 × 10⁻²) significantly improved mean SAV compared to OFF. KaDBS did not reach significance compared to OFF (p=0.115; table S1).

DBS condition showed no significant effect on arrhythmicity (F(3,18.9)=2.934, p=0.060, ηp²=0.32, n=6; table S1). Two baseline freezers (Participants 02 and 06) had unmeasurable arrhythmicity values during OFF due to complete freezing, reducing the effective sample size from 8 to 6 for the OFF condition. Participants demonstrated an arrhythmicity of 117.77 ± 32.61 (estimated marginal mean ± SE) during OFF DBS, with all DBS conditions reducing gait arrhythmicity: cDBS by 68.5 ± 27.0, KaDBS by 59.9 ± 27.0, and iDBS by 72.5 ± 27.0. The lack of data from the two participants who were frozen throughout the stepping-in-place trial likely diluted the significance of the overall effect of DBS condition on gait arrhythmicity, as these individuals demonstrated substantial arrhythmicity reductions when ON DBS.

### Kinematic Adaptive DBS Improved Gait in Free Walking Task

From the cohort of participants with GI&FOG (n=8) from the SIP task, seven participants (n=7) completed the TBC task. Participant 06 did not perform the TBC course due to inability to safely perform the task without assistance under cDBS. The TBC consisted of two walking patterns: elliptical and figure-of-eight courses. Each participant performed both walking patterns under each stimulation condition, with each pattern analyzed as a separate trial. Percent time freezing showed a significant effect of DBS condition (F(3,45)=6.94, p = 6.17 × 10⁻⁴, ηp²=0.32; Supplementary Table 1). During OFF stimulation, participants spent 40.36 ± 10.19% (estimated marginal mean ± SE) of the time freezing. All DBS conditions showed reductions in freezing: KaDBS by 33.40 ± 12.74% (p = 9.00 × 10⁻⁴), cDBS by 31.74 ± 12.74% (p = 1.00 × 10⁻³), and iDBS by 27.13 ± 12.74% (p = 8.8 × 10⁻³), (Fig. 6A; table S1).

**Fig. 6.**
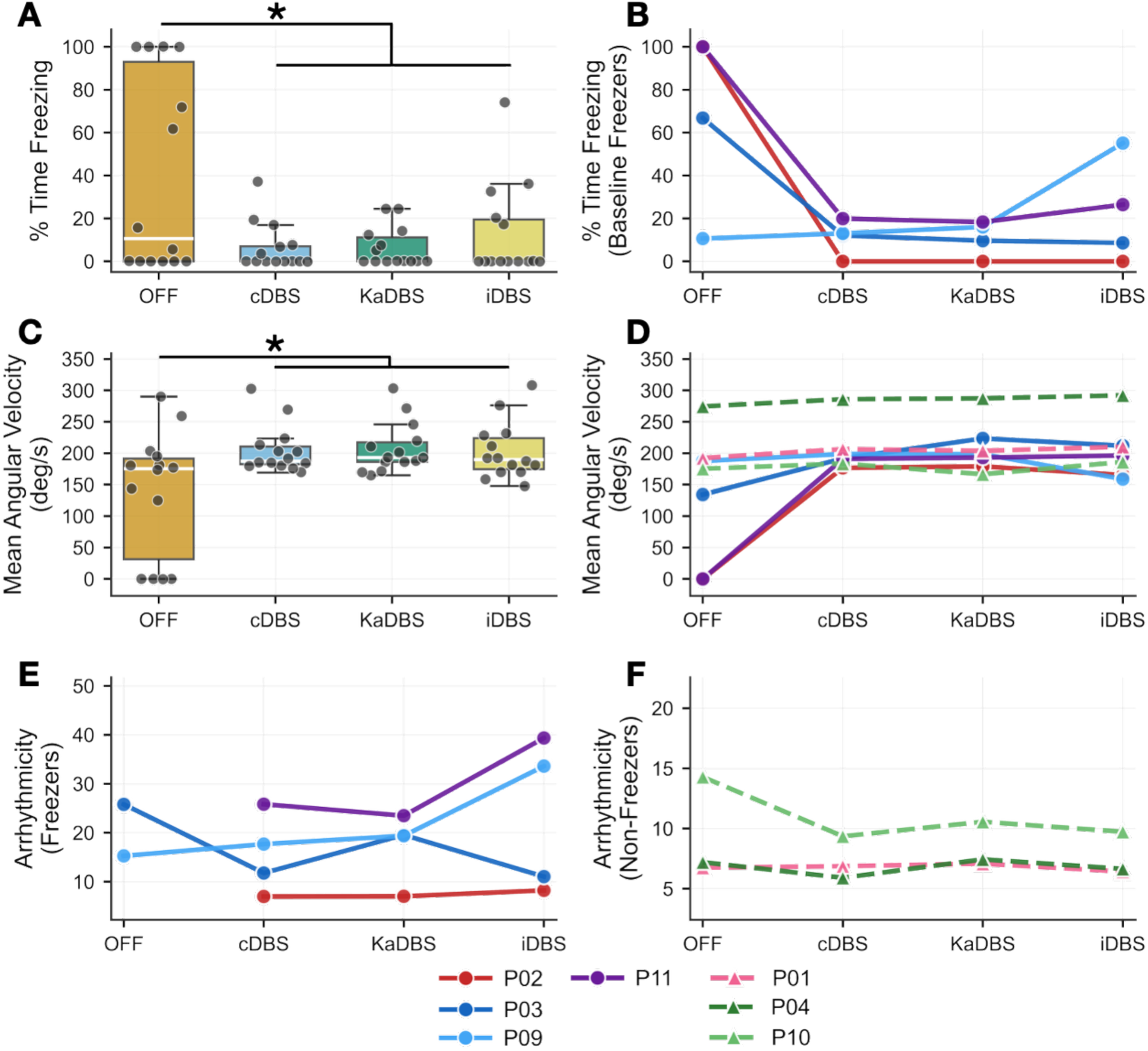
Gait metrics across stimulation conditions during turning-and-barrier course. (**A**) Population % time freezing (N=7) shown as colored boxplots with individual data points along with mean and standard deviation. * Indicates p < 0.05. (**B**) % Time freezing trends for baseline freezers. (**C**) Population mean shank angular velocity (N=7) shown as colored boxplots with individual data points along with mean and standard deviation. * Indicates p < 0.05. (**D**) Mean shank angular velocity trends for baseline freezers and for non-freezers. Arrhythmicity trends for (**E**) baseline freezers and (**F**) for non-freezers. Baseline freezers (solid lines, circles, n=4) defined as participants with >0% freezing during OFF condition, non-freezers (dashed lines, triangles, n=3) defined as participants with 0% baseline freezing. Turning-and-barrier course included elliptical and figure-eight walking patterns. In boxplots (**A**, **C**), each participant contributes two data points per stimulation condition representing each task type. In trajectory plots (**B**, **D**, **E**, **F**), values are averaged across both task types for each participant per condition.

Participants were stratified based on baseline freezing behavior: baseline freezers exhibited >0% freezing during OFF stimulation (n=4), while non-freezers demonstrated 0% freezing during OFF stimulation (n=3). Individual trends (averaged across both task types) revealed heterogeneous treatment effects among baseline freezers (Fig. 6B). Complete elimination of % time freezing was achieved by Participant 02 across all DBS conditions (100% OFF to 0%). Participant 11 showed substantial improvement from complete freezing (100% OFF) with optimal responses observed during KaDBS (19.15%) compared to cDBS (26.63%) and iDBS (28.43%). Participant 03 demonstrated better performance with KaDBS (9.54%) compared to cDBS (13.61%) and iDBS (10.23%). To explore the relationship between therapeutic benefit and total stimulation dose, we examined the total electrical energy delivered (TEED (1s)) in this participant. Equivalent energy delivery across conditions (78 μW) was associated with differential freezing outcomes, with KaDBS having the lowest freezing (fig. S2). Conversely, Participant 09 had increased freezing with all DBS conditions compared to OFF (13.31%), with particularly poor performance during iDBS (53.95%) while experiencing moderate freezing during KaDBS (20.02%) and cDBS (20.10%). Non-freezers (Participants 01, 04, 10) maintained 0% freezing across all DBS conditions (table S2).

Mean shank angular velocity showed a significant effect of DBS condition (F(3,45)=10.37, p = 2.63 × 10⁻⁵, ηp²=0.41; table S1). During OFF stimulation, participants had a mean SAV of 136.41 ± 23.40 deg/s (estimated marginal mean ± SE), with all DBS conditions showing improvements: KaDBS by 68.25 ± 22.80 deg/s (p = 1.00 × 10⁻⁴), cDBS by 64.71 ± 22.80 deg/s (p = 1.00 × 10⁻⁴), and iDBS by 63.08 ± 22.80 deg/s (p = 2.00 × 10⁻⁴), (Fig. 6C; table S1).

With only 5 participants providing OFF condition data due to complete freezing in participants 02 and 11, statistical analysis was underpowered. During OFF stimulation, participants had an arrhythmicity of 16.20 ± 3.22 (estimated marginal mean ± SE), with minimal changes observed across stimulation conditions: cDBS reduced arrhythmicity by 2.94 ± 3.22, KaDBS by 1.62 ± 3.22, and iDBS increased by 1.15 ± 3.22. However, lack of data from the two participants who were frozen throughout the free walking trial likely masked the effect of DBS condition on arrhythmicity, as these individuals were able to perform the task when ON DBS.

## Discussion

This first clinical trial of KaDBS for PD demonstrated that real-time kinematic control of DBS is safe, well-tolerated, and effective for reducing gait impairment and freezing of gait. Both KaDBS algorithms demonstrated favorable safety profiles compared to clinical cDBS. The arrhythmicity model achieved the highest proportion of symptom-free reports (87.5%), followed by the P(FOG) model (71.4%) and cDBS (50.0%). Adverse sensations were limited to mild, transient imbalance and dizziness during KaDBS, whereas cDBS produced a broader range of symptoms including pulling and nausea (Fig. 4, A to C). Across the cohort, KaDBS significantly reduced percent time freezing compared to OFF stimulation in both the harnessed stepping-in-place task (35.8 ± 9.9% reduction, p = 4.80 × 10⁻³, Fig. 5A, table S1) and the naturalistic turning and barrier course (33.4 ± 12.7% reduction, p = 9.00 × 10⁻⁴, Fig. 6A, table S1). These findings represent the first demonstration that adaptive neuromodulation can effectively target the episodic nature of gait dysfunction in PD using behavioral control signals.

Stratification by baseline freezing phenotype revealed that therapeutic effects were concentrated in participants who exhibited freezing during OFF stimulation. Among baseline freezers (n=5 for SIP, n=4 for TBC), DBS conditions produced substantial reductions in freezing episodes (Fig. 5B and 6B), while non-freezers (n=3 for both tasks) maintained normal gait function across all conditions (table S2). This pattern confirms that KaDBS can precisely target pathological motor states without disrupting normal motor control, an essential property for any adaptive neuromodulation system intended for continuous use during activities of daily living.

Individual response trends demonstrated clinically meaningful heterogeneity that warrants careful consideration for future implementations. During SIP, Participant 03 achieved complete freezing elimination selectively with KaDBS (from 76.09% OFF to 0% ON), while retaining residual episodes with cDBS (7.38%) and iDBS (14.93%) (Fig. 5B). This selective response persisted during the more challenging TBC task, where the same participant maintained better performance with KaDBS (9.54%) compared to cDBS (13.61%) and iDBS (10.23%) (Fig. 6B). Participant 11 showed substantial improvement from complete freezing during TBC (100% OFF) with optimal responses observed during KaDBS (19.15%) compared to cDBS (26.63%) and iDBS (28.43%) (Fig. 6B). These algorithm-specific benefits suggest that kinematic control strategies may provide advantages over conventional cDBS for certain individuals, potentially reflecting differences in how gait networks respond to stimulus timing. Analysis of TEED in Participant 03 revealed that therapeutic benefit was not attributable to reduced energy dose (fig. S2). KaDBS achieved better freezing outcomes despite delivering equivalent total energy compared to cDBS and iDBS, suggesting that temporal alignment of stimulation with real-time kinematic biomarkers, rather than cumulative energy delivery, underlies the observed therapeutic differences. However, other participants showed equivalent or superior responses to cDBS. Participant 04 demonstrated lower freezing with cDBS (0%) compared to KaDBS (10.80%) during SIP (Fig. 5B), though all DBS conditions eliminated freezing during TBC (table S2). Participant 02 had complete elimination of freezing across all DBS conditions in both tasks (Fig. 5B and 6B). This variability highlights a fundamental challenge in personalized neuromodulation: not all patients will respond optimally to the same control strategy. The existence of algorithm-specific responders argues for systematic methods to identify which patients are most likely to benefit from adaptive versus conventional continuous approaches.

The comparable performance between KaDBS and cDBS during acute controlled assessments reflects our experimental design and study objectives. We sought to establish feasibility, safety, and efficacy for reducing freezing rather than demonstrate superiority over cDBS in brief laboratory trials. All DBS conditions operated within clinically established therapeutic windows (Imin to Imax) as established during calibration (Fig. 2), where even minimum stimulation (Imin) provided meaningful clinical benefit. The short-duration, controlled assessment likely favored cDBS, as participants had recently undergone clinical optimization of their stimulation parameters by a movement disorder specialist prior to assessments. This recent optimization likely maximized cDBS performance during our acute assessments.

The potential advantages of KaDBS, including dynamic response to fluctuating symptoms and reduction of unnecessary stimulation exposure, may emerge most clearly during extended at-home monitoring where symptom expression varies across medication states, environmental contexts, and daily activities. This developmental trajectory parallels neural aDBS, where early investigations including the ADAPT-PD trial employed non-inferiority designs demonstrating that aDBS was tolerable and as effective as cDBS(*15*), while subsequent chronic ambulatory studies revealed differential benefits during extended at-home use where symptom fluctuations demanded adaptive responsiveness(*18, 34*).

The unexpected comparable performance of iDBS merits careful evaluation in the context of our testing protocol. Despite lacking physiological timing for its adjustments, the iDBS condition had significant improvements in gait metrics comparable to both cDBS and KaDBS (Fig. 5 to 6). This suggests that within short testing periods, stochastic variation of stimulation may have been sufficient to disrupt pathological basal ganglia dynamics regardless of timing precision. An alternative explanation is that our therapeutic windows were sufficiently well-calibrated that even random adjustments remained within a beneficial range. The equivalence observed here likely reflects our controlled testing paradigm.

Task-dependent dissociations in treatment response showed important limitations of current approaches. Participant 06 demonstrated modest improvement across all DBS conditions during SIP (Fig. 5B, table S2) but could not safely complete the TBC while on cDBS, requiring study discontinuation. Participant 09 showed no freezing during SIP across all conditions (Fig. 5B, table S2) but exhibited paradoxical worsening with all active DBS conditions during TBC (OFF: 13.31%, cDBS: 20.10%, KaDBS: 20.02%, iDBS: 53.95%) (Fig. 6B, table S2). Given the temporal separation between the TBC and SIP tasks (approximately 3 months) and the difference between harnessed controlled SIP versus free ambulatory walking with turns and obstacles, this pattern likely reflects both disease progression and the inherent challenges of complex locomotor environments. Examination of KaDBS parameters reveals that while stimulation amplitude ranges were expanded, ramp-up rates were reduced from 2.4/1.6 mA/s during SIP to 0.25/0.25 mA/s during TBC (table 2). Combined with the algorithm switch from arrhythmicity-based to probabilistic FOG detection, this created a system that may have detected freezing episodes but responded too slowly to provide therapeutic intervention before episodes resolved spontaneously. The poor performance of all DBS modalities for this participant, including optimized cDBS, suggests that stimulation parameters effective for controlled stepping may be counterproductive during challenging ambulatory tasks that require greater motor flexibility and adaptive responses.

Statistical non-significance for arrhythmicity metrics reflected measurement constraints rather than therapeutic inefficacy. Participants who were completely frozen during OFF testing (n=2 for SIP, n=2 for TBC) had no detectable steps, precluding arrhythmicity calculation. This systematically excluded the most severely impaired participants from analysis, biasing group statistics toward null findings. Among participants with measurable OFF values, individual patterns consistently showed arrhythmicity reductions with DBS. Participant 03 transitioned from severely arrhythmic stepping (227.42) to near-normal rhythmicity with KaDBS (23.36) during SIP. Participants 02 and 06, who were frozen throughout SIP-OFF, achieved functional stepping rhythmicity when ON DBS. These trends confirm that gait arrhythmicity improvements occurred but were masked by missing data from the most responsive individuals.

The stimulation adjustment “ramp rates” utilized during KaDBS in this study represent a meaningful advance over neural aDBS approaches. Four of eight participants tolerated rapid stimulation adjustments (up to 4.0 mA/s), faster than rates typically used in neural aDBS (max around 0.1 mA/s)(*14, 15, 17, 35–38*). This rapid responsiveness allows the system to react to sudden freezing episodes with minimal delay, matching the temporal characteristics of the symptom itself. The variation in tolerated ramp rates between participants (table 2) highlights the importance of individualized calibration protocol, as we have detailed in our methods, for adaptive stimulation protocols. Importantly, we did not observe a clear relationship between disease severity and rapid ramp rate tolerability, suggesting that factors beyond symptom severity likely influence individual tolerance to stimulation adjustments(*39*).

KaDBS demonstrates practical advantages over neural aDBS approaches. Kinematic signal acquisition avoids stimulation artifacts and sensing configuration constraints that complicate neural recordings(*40*). This enabled continued use of clinically optimized stimulation parameters without sensing compromises typically required in neural aDBS. Kinematic approaches have shown therapeutic promise for other movement disorders including tremor(*31, 41*), suggesting broader applications for peripheral sensing in adaptive neuromodulation. Kinematic sensing provides direct motor symptom measurement rather than neural surrogates. While subthalamic beta oscillations exhibit inter-participant variability and depend on electrode placement precision (*17*), kinematic signatures of GI&FOG appear to show more consistent patterns across participants (*42*). Despite heterogeneity in disease duration (12-19 years) and symptom profiles (table 1), participants demonstrated similar kinematic responses to therapeutic DBS, though a larger sample would be needed to confirm generalizability across PD subtypes.

Future developments should prioritize hybrid control systems integrating kinematic and neural biomarkers(*43*). Combined approaches could provide complementary information during different behavioral states. Recent advances demonstrate that ensemble algorithms significantly improve FOG detection accuracy and reveal temporal patterns in symptom presentation(*44*). FOG episodes exhibit circadian rhythmicity, suggesting potential benefits from time-dependent intervention strategies(*44*). Advanced machine learning architectures could integrate temporal dependencies with real-time kinematic data, enabling context-aware adaptive systems for personalized neuromodulation. Substantial miniaturization and sensor integration advances remain necessary for ambulatory implementation. Expanding commercial wearable technology availability supports such developments (*30*). Future investigations should evaluate KaDBS benefits for other motor symptoms and assess long-term efficacy and medication requirement impacts.

Several limitations require acknowledgment. Despite being the largest KaDBS cohort to date, our sample size of eight participants limits broad generalizability and statistical power. Our “computer-in-the-loop” implementation, while necessary for this initial investigation, introduces complexity incompatible with routine clinical use. The system’s current inability to distinguish between intentional stopping and pathological freezing creates potential for inappropriate stimulation increases during normal stops, limiting application in activities of daily living. This was not the goal of this initial feasibility study but is relevant for future advancement. This challenge parallels difficulties in neural aDBS where context-specific interpretation of neural signals remains challenging(*45*). Also, the narrow therapeutic windows employed for safety, while necessary, may have limited our ability to detect superiority of KaDBS over conventional cDBS approaches. Assessment periods were brief (100 seconds for SIP, approximately 200-300 seconds per TBC trial), favoring recently optimized cDBS over adaptive strategies whose benefits likely emerge during extended use with fluctuating symptoms.

The reliance on external sensors presents challenges for long-term use, including mounting inconsistencies, signal drift, signal dropouts, and battery limitations. The calibration procedures required to establish personalized therapeutic windows, ramp rates, and detection thresholds represent a significant burden that would need streamlining and automation for widespread adoption. Embedded accelerometers in next-generation implantable pulse generators could eliminate the need for external hardware, while automated calibration algorithms could reduce the clinical burden of individualized parameter selection. Transitioning from the current computer-in-the-loop architecture to fully implantable systems that integrate kinematic and neural biomarkers would enable the chronic ambulatory use where the advantages of adaptive over continuous stimulation for episodic symptoms like freezing of gait are most likely to emerge.

## Materials and methods

### Study Design

This was a single-center exploratory clinical trial evaluating the safety, tolerability, and efficacy of KaDBS for GI&FOG in PD (ClinicalTrials.gov: NCT04043403). The trial used a within-subjects crossover design in which each participant was assessed under four randomized, double-blind stimulation conditions: OFF, cDBS, KaDBS, and iDBS. The study was designed as an exploratory feasibility investigation rather than a superiority or non-inferiority trial. The primary outcome was safety and tolerability, assessed through participant-reported symptoms collected after each stimulation condition. Secondary outcomes were percent time freezing, gait arrhythmicity, and mean shank angular velocity, measured during both the SIP and TBC tasks. The initial target enrollment was 15 participants; 8 were enrolled before recruitment was halted due to the discontinuation of the Summit RC+S by the manufacturer. The study protocol was approved by Stanford University’s Institutional Review Board (IRB-52548) and supported by an Investigational Device Exemption from the U.S. Food and Drug Administration.

### Participants

Eligibility criteria required participants to be at least 18 years old, with complications from medication (e.g., dyskinesia, medication refractory tremor, or fluctuations in symptom response), and noticeable impact on quality of life, regardless of medication status. Additionally, participants needed to score at least 1 on the freezing of gait questionnaire (FOG-Q) and/or the gait-related item (3.10) of the Movement Disorders Society Unified Parkinson’s Disease Rating Scale Part III (MDS-UPDRS III). All participants were implanted with bilateral DBS leads (model 3389) targeting the sensorimotor region of the STN and the Summit RC+S investigative sensing neurostimulator (Medtronic, Inc)(*46*).

Exclusion criteria included being over 80 years old, having dementia, untreated psychiatric conditions, Hoehn and Yahr stage 5 classification, any significant surgical risks, or contraindications such as cardiac pacemakers, a need for rTMS, ECT, MRI, diathermy, pregnancy, cranial metallic implants, or a history of seizures. Participants provided written consent.

### Experimental Protocol

All participants were clinically optimized on their DBS parameters prior to experimental assessments. We conducted a double-blind investigation comprising two visits separated by three months (3 to 6 testing days each), following a bench-to-bedside progression (fig. S1). All assessments occurred in the practically defined off-medication state, requiring withdrawal from long-acting dopamine agonists (at least 48 hours), controlled-release carbidopa/levodopa (at least 24 hours), and immediate-release medications (at least 12 hours). For active DBS conditions, testing was performed after a 20-minute wash-in period. For OFF, stimulation was turned off for at least 15 minutes prior to the gait task to allow washout. The iDBS control condition was configured to match the KaDBS therapeutic window, ramp rates, and temporal dynamics but with stochastic amplitude variation independent of the participant’s gait state. All active conditions used identical contact configurations, frequency (140.1 Hz), and pulse width (60 μs).

Visit 1 evaluated safety and tolerability using the validated 100-second SIP task performed on force-instrumented platforms with participants safely harnessed(*47*). Following successful completion, visit 2 employed the validated TBC task designed to elicit freezing of gait during naturalistic free walking(*42*). The TBC task incorporated dividers simulating real-world environments including narrow hallways and doorways. Participants completed two elliptical walking patterns followed by two figure-eight patterns.

Prior to evaluating the four stimulation conditions, we performed calibration to determine optimal KaDBS parameters for each participant. The therapeutic window defines the permissible DBS amplitude adaptation range (Imax – Imin) for KaDBS, ensuring stimulation adjustments remain within amplitudes that provide clinical benefit while avoiding side effects. To establish therapeutic windows, we performed randomized stimulation titrations spanning 25-125% of participants’ clinical stimulation amplitudes. Each participant also underwent systematic ramp rate testing to determine tolerable rates of stimulation adjustment and based on individual tolerability we selected the appropriate stimulation control mode (see Control Algorithm Implementation). The calibration phase concluded with short validation runs of the established parameters during the gait task for that visit. Individual calibration parameters are summarized in table 2.

### Real-Time Gait Feature Extraction and Freezing Classification

The gyroscope data were streamed from both bilateral shank IMU sensors at 128 Hz. The data processing pipeline developed from previous work performed five key operations in real-time(*32*). First, signal preprocessing applied a first-order 9 Hz low-pass filter to preserve step features while removing noise:

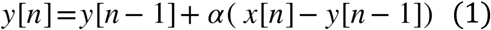

where x[n] is the raw signal sample, y[n] is the filtered output, and α=0.306416 to reflect a first order 9 Hz low-pass filter.

Second, step detection identified peaks exceeding 10°/s with minimum temporal separation of 400ms for turning-based tasks and 600ms for stepping-in-place tasks. These constraints were selected to match typical human step cadence while eliminating non gait movements (*42, 48*).

Third, gait features were computed from each detected step. Swing angular range (SAR) was calculated as the summation of positive angular (ω) between zero crossings:

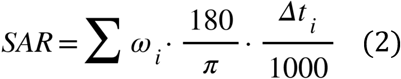

where ω_i_ is angular velocity in rad/s at sample i, and Δt_i_ is the time difference in milliseconds between samples. Stride time (ST) was defined as the interval between consecutive peaks. Asymmetry (AS) was calculated from left and right swing times (ST_left_, ST_right_) using the last 3 swing times per leg:

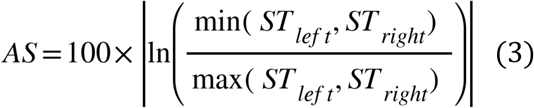

Fourth, gait arrhythmicity (AR_i_) was calculated as the average coefficient of variation (CV = σ/μ, where σ and μ are standard deviation and mean of stride times, respectively) from both legs using the last 3 stride times per leg:

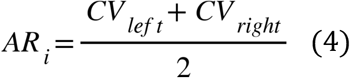

Finally, freezing probability (P_i_) was estimated using a validated logistic regression model(*42*):

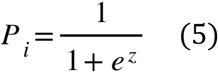

where z = – (0.941 + 2.034 *AR_i_* + 0.0931 *ST* – 0.0615 *SAR* + 0.0003 *AS*)

These features were computed continuously on a rolling window of the most recent detected steps, with stimulation decisions updated at a 1Hz rate to balance responsiveness with system stability and provide a stimulation decision on a step-by-step basis. Technical challenges encountered during implementation, including maintaining stimulation control during akinetic freezing episodes and managing communication latency between system components, are detailed in the Supplementary Methods.

### Control Algorithm Implementation

Both the arrhythmicity threshold and FOG classifier control policies utilized one of two stimulation adjustment mechanisms, selected based on individual participant tolerance. For participants who tolerated rapid stimulation changes (>0.1 we implemented a Go-To-States mode that employed a state table containing up to nine predefined states. Each state specified independent control of bilateral STN stimulation (increase, decrease, or hold), allowing various combinations such as bilateral increases, unilateral adjustments, or opposing changes between STNs, with defined stimulation amplitudes for each state. Upon receiving a decision from either control algorithm, the system selected a state and initiated stimulation changes with 200-300 milliseconds command latency. Stimulation amplitude was adjusted at the specified ramp rate until reaching the target amplitude or receiving a new state command.

For participants requiring slower stimulation adjustments (<0.1 mA/sec), we implemented an Increments/Decrements (Inc/Dec) mode that modified stimulation in discrete 0.1 mA steps. This method addressed a timing conflict in the Go-To-States mode at slower ramp rates. Since our control system updated at 1 Hz but a stimulation change at <0.1 mA/sec would require >1 second to complete, each new command would override the previous command before implementation. The Inc/Dec mode ensured that each adjustment was completed within the update cycle, preventing command cancellation and allowing sequential execution of control algorithm decisions. Both modes operated through a UDP interface, enabling dynamic parameter adjustment within predefined therapeutic windows based on the control algorithms’ real-time decisions.

The control algorithm implemented asymmetric ramp rates: amplitude increases occurred faster than decreases to prevent rapid stimulation withdrawal following FOG resolution, which could precipitate symptom recurrence.

### Data acquisition and offline data Analysis

#### Stepping-in-Place (SIP) Analysis

Kinetic data were obtained through vertical ground reaction forces, which were normalized to participant body weight and parsed into discrete gait cycles (detailed protocol in Supplementary Materials). Temporal parameters including swing times and stride times were extracted to quantify gait arrhythmicity. Arrhythmicity was determined by calculating the CV of the stride time averaged across both legs, with higher CV values indicating increased arrhythmicity. Freezing of gait (FOG) episodes were identified within the force plate data using a validated algorithmic approach(*47*). For Participant 10, where force plate data were corrupted, FOG detection utilized kinematic parameters derived from IMUs, following a validated logistic regression approach(*48*). The percentage of time spent freezing was then calculated as the total duration of FOG episodes divided by the overall trial time (i.e. 100 seconds).

#### Turning and Barrier Course (TBC) Analysis

As the TBC consisted of two subtasks: walking in ellipses and then in figures-of-eight, the task performance for each subtask were analyzed separately. Triaxial angular velocity data from shank IMUs underwent preprocessing using a zero-phase 8th order Butterworth low-pass filter (9 Hz cut-off frequency). Principal component analysis was implemented to extract 1-D shank angular velocity (SAV) in the sagittal plane (*42*), which were subsequently utilized for gait cycle segmentation (protocol detailed in Supplementary Materials). Arrhythmicity metric was computed using the same methodology as the stepping-in-place analysis. FOG episodes were detected using a validated logistic regression model that incorporated multiple gait parameters: arrhythmicity and asymmetry over the last six steps, stride time and swing angular range from the last step(*42*). FOG events were defined when the model’s computed freeze probability exceeded a threshold of 0.7. The percentage of time spent freezing was then calculated as the total duration of FOG episodes divided by the total trial time.

#### Statistical Analysis

Statistical analyses were performed using R (version 3.6.0) with lme4 (v1.1-31), lmerTest (v3.1-3), emmeans (v1.8.8), and effectsize (v0.8.3) packages. For each outcome measure (freezing, arrhythmicity, and mean shank angular velocity), linear mixed effects models were fitted with participant as a random intercept. For SIP analyses, models included stimulation condition (OFF, cDBS, aDBS, iDBS) as a fixed effect (model formula: outcome ∼ condition + (1│participant)), while for TBC analyses, models included both stimulation condition and task type (elliptical and figure-eight patterns) as fixed effects (model formula: outcome ∼ condition + task + (1│participant)). All multiple comparisons were adjusted using Sidak’s method, with adjusted p < 0.05 indicating statistical significance. When main effects were significant, post-hoc pairwise comparisons of each DBS condition against baseline (OFF) were performed using estimated marginal means with the same Sidak’s adjustment. Effect sizes were quantified using partial eta squared (np^2^) with 95% confidence intervals. Results are presented as estimated marginal means ± standard error.

## Data Availability

All data needed to evaluate the conclusions in the paper are present in the paper and/or the supplementary materials. Additional data related to this paper may be requested from the corresponding author.

## Acknowledgments

The authors would like to thank the participants who dedicated their time to this study and Medtronic, Inc for providing the devices without additional financial support.

## Funding

This work was supported by:

National Institute of Neurological Disorders and Stroke grant UH3NS107709 (HMBS)

National Institute of Neurological Disorders and Stroke grant UH3NS128150 (HMBS)

National Institute of Neurological Disorders and Stroke U24NS113637 (HMBS)

Robert and Ruth Halperin Foundation (HMBS)

John A. Blume Foundation (HMBS)

John E Cahill Family Foundation (HMBS)

## Author Contributions

Conceptualization: Y.M.K., M.N.P., K.B.W., J.O., J.A.H., and H.M.B.S.

Methodology: S.K., Y.M.K., M.N.P., L.P., P.A.¹, K.B.W., J.O., H.J.D., and J.A.H.

Investigation: S.K., Y.M.K., M.N.P., L.P., P.A.¹, P.A.², E.F.L., J.A.M., K.B.W., J.O., H.J.D., C.D., A.S.G., C.C., and S.L.H.

Visualization: S.K., Y.M.K., M.N.P., K.B.W., H.J.D., C.C., and P.A.²

Funding acquisition: H.M.B.S.

Project administration: M.N.P., L.P., P.A.¹, E.F.L., J.A.M., K.B.W., H.J.D., and S.L.H.; Supervision: M.N.P., K.B.W., J.A.H., and H.M.B.S.

Writing—original draft: S.K. and Y.M.K.

Writing—review and editing: S.K., Y.M.K., M.N.P., L.P., P.A.¹, E.F.L., J.A.M., K.B.W.,

J.O., H.J.D., C.D., A.S.G., C.C., S.L.H., P.A.², J.A.H., and H.M.B.S.

¹P.A. refers to Pranav Akella; ²P.A. refers to Prerana Acharyya.

## Supplementary Materials and Methods

### Technical Challenges in the Distributed Architecture

#### Control policy for abrupt freezes

During system development and initial data collection, we identified and addressed two critical technical challenges in the distributed architecture. The first challenge emerged when participants experienced akinetic freezing episodes. In most scenarios, freezing episodes are accompanied by step deterioration prior to complete akinesia, which our main control algorithm can detect through its continuous processing loop analyzing the last six steps to capture increasing freezing probability and determine stimulation adjustments. However, when participants transitioned into freezing abruptly following generally normal steps, the algorithm would make decisions based solely on the last six steps in the buffer, which remained unchanged during the freeze (as no new steps are detected). If these steps happened to have characteristics of normal gait (low ARi or Pi), the system would incorrectly classify the state as non-freezing despite the actual occurrence of a freezing episode, potentially delaying appropriate stimulation increases.

To address this scenario, we implemented an additional algorithm that tracks the consistency of calculated arrhythmicity (ARi) and freezing probability (Pi) values across consecutive processing cycles. The system stores these values from each decision point and compares them to subsequent calculations. During normal stepping, these values naturally vary as new step data enters the buffer. However, during a freezing episode, these values remain identical across consecutive processing cycles since no new steps are being detected to replace the data in the buffer. If the system detects identical ARi and Pi values for two consecutive processing cycles (approximately 2 seconds, selected as a balance between responsiveness and false positive prevention), it identifies this as a freezing state regardless of the absolute values in the step buffer, and responds accordingly. These decisions are then executed via either the Go-To-State mechanism (immediately transitioning to predefined stimulation states, including Imax when freezing is detected) or the Increments/Decrements mechanism (gradually adjusting in 0.1 mA steps for participants requiring slower transitions). This freezing detection continues until new steps are identified, causing changes in the computed ARi/Pi values, indicating altered gait state.

#### Fall back mode for sensor wireless communication latency

The second challenge involved managing communication latency between components of the distributed system architecture. We observed occasional interruptions in data flow between the APDM sensors, processing laptop, and the Summit neurostimulator API. During periods of connection loss, the APDM sensors buffer incoming kinematic data locally until connection is reestablished, but the gait processing application cannot analyze this data in real-time. To detect these interruptions, we implemented a latency monitoring system that continuously tracks the time difference between consecutive data batches received from each sensor. If this difference exceeds 500 milliseconds (indicating potential data buffering issues where batches arrive too quickly as the connection is reestablished), the system enters a predefined contingency mode where stimulation amplitude is held at current levels to maintain stability. When normal communication is restored, the system processes only the most recent six steps from the buffered data, typically introducing a brief 1-2 second lag during system resynchronization before resuming normal adaptive control.

#### Kinetic and Kinematic Data Analysis

Stepping-in-Place: For each limb, stride time was defined as the interval from initial ground contact to the next contact of the same foot. Swing time was the interval from toe-off (foot leaving the force plate) to the subsequent contact of that foot on the plate. From these two measures, we derived asymmetry and arrhythmicity using standard formulations based on inter-limb differences and stride-to-stride variability, respectively. Kinetic data was obtained from the dual force plates (Bertec Corporation, Columbus, OH, USA), sampled at 1000 Hz.

Turning and Barrier Course: Lower-limb kinematics were quantified from shank angular velocity in the sagittal plane. The onset of swing was identified at the zero-crossing where angular velocity transitioned from negative to positive (rising through zero), and the end of swing at the next zero-crossing in the opposite direction (falling through zero). Peak shank angular velocity for each step was taken as the first positive local maximum following swing onset.

Two temporal metrics were computed: (i) forward swing duration, defined as the time between consecutive zero-crossings for the same limb that bound the swing phase, and (ii) stride time, defined as the interval between successive peak angular velocities of the same limb. We used peaks in shank angular velocity to delineate steps (an approach that circumvents ambiguity in heel-strike detection in Parkinson’s disease)(*1*) retaining only peaks that exceeded a minimum amplitude of 10°·s⁻¹ for the Turning and Barrier Course. The swing angular range was obtained by integrating the sagittal angular velocity over each swing phase.

#### TEED Analysis

Total electrical energy delivered per second (TEED (1s)) was calculated using: TEED (1s) = I² × pw × f × R, where I = current (A), pw = pulse width (s), f = frequency (Hz), and R = impedance (Ω)(*2*). For double monopolar configurations, TEED was calculated separately for each active contact and summed. TEED was computed at one-second intervals and averaged across each trial.

## Supplementary Results

**Fig. S1.**
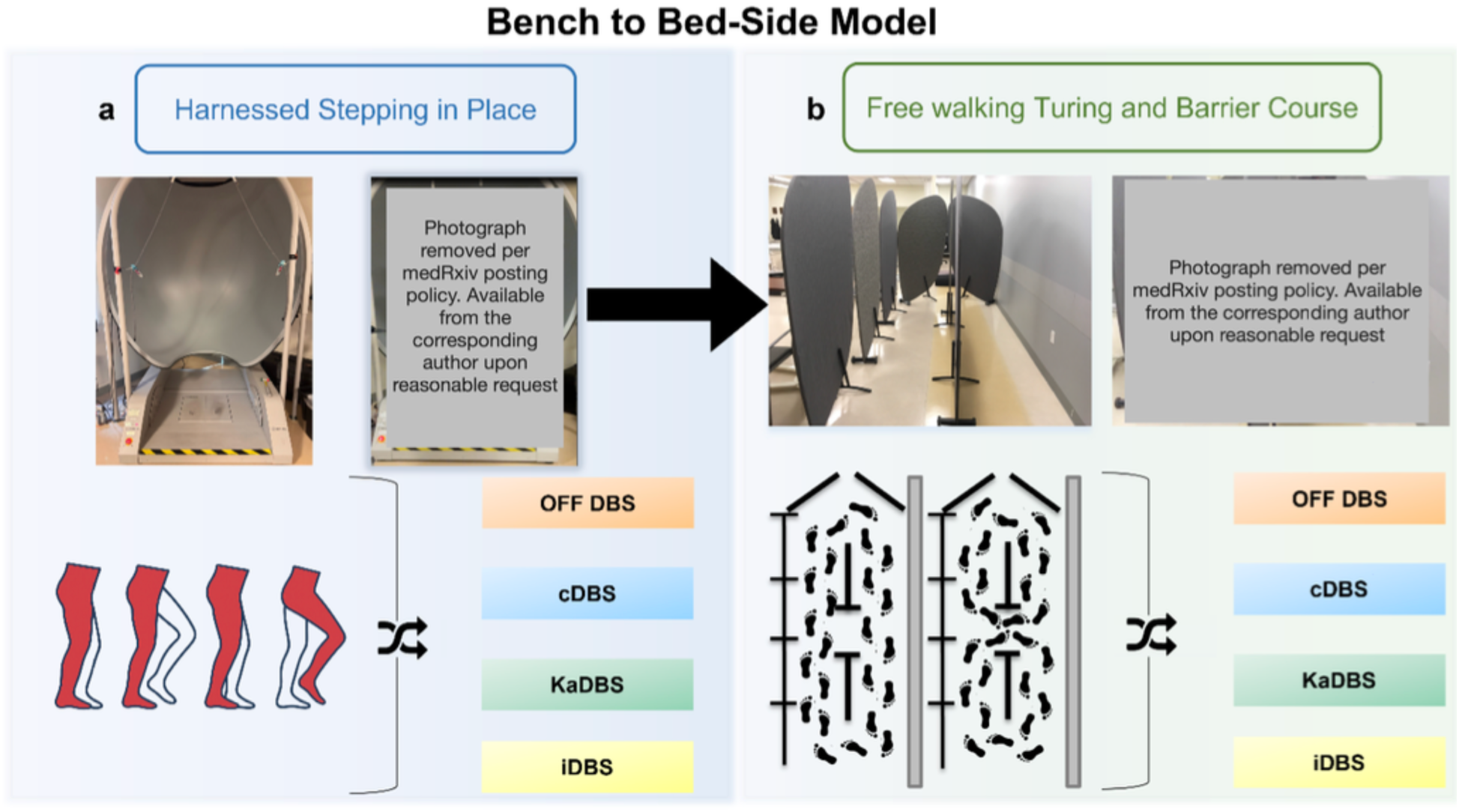
Study design for kinematic adaptive deep brain stimulation (KaDBS) evaluation. A progressive two-phase protocol from controlled to naturalistic environments. **a,** Harnessed stepping-in-place task (Visit 1) used for initial safety and efficacy evaluation, with participants secured in a safety harness while stepping on force plates. **b,** Free walking turning and barrier course (Visit 2) with narrow passages designed to elicit freezing of gait. Both phases tested four stimulation conditions (OFF DBS, cDBS, KaDBS, iDBS) in randomized order. Only participants completing Visit 1 advanced to Visit 2. cDBS, continuous deep brain stimulation; iDBS, random intermittent deep brain stimulation.

**Supplementary Table 1:** Linear Mixed-Effects Model Analysis of DBS Effects on Gait Parameters.

| Metric & Comparison | Test Statistic | df | P value | Effect Size |
| --- | --- | --- | --- | --- |
| *Percent Time Freezing* |  |  |  |  |
| *Stepping-in-Place Task* |  |  |  |  |
| Condition main effect | $F_{3,21} = 5.985$ | 3, 21 | $4.10 \times 10^{-3}$ | $\eta_p^2 = 0.46 [0.14, 1.00]$ |
| OFF vs KaDBS | $t_{21} = 3.618$ | 21 | $4.80 \times 10^{-3}$ | — |
| OFF vs cDBS | $t_{21} = 3.416$ | 21 | $7.80 \times 10^{-3}$ | — |
| OFF vs iDBS | $t_{21} = 3.319$ | 21 | $9.80 \times 10^{-3}$ | — |
| *Turning and Barrier Course* |  |  |  |  |
| Condition main effect | $F_{3,45} = 6.936$ | 3, 45 | $6.17 \times 10^{-4}$ | $\eta_p^2 = 0.32 [0.11, 1.00]$ |
| OFF vs KaDBS | $t_{45} = 3.918$ | 45 | $9.00 \times 10^{-4}$ | — |
| OFF vs cDBS | $t_{45} = 3.901$ | 45 | $1.00 \times 10^{-3}$ | — |
| OFF vs iDBS | $t_{45} = 3.143$ | 45 | $8.8 \times 10^{-3}$ | — |
| *Shank Angular Velocity* |  |  |  |  |
| *Stepping-in-Place Task* |  |  |  |  |
| Condition main effect | $F_{3,21} = 3.504$ | 3, 21 | $3.33 \times 10^{-2}$ | $\eta_p^2 = 0.33 [0.02, 1.00]$ |
| OFF vs iDBS | $t_{21} = 2.906$ | 21 | $2.51 \times 10^{-2}$ | — |
| OFF vs cDBS | $t_{21} = 2.645$ | 21 | $4.48 \times 10^{-2}$ | — |
| OFF vs KaDBS | $t_{21} = 2.189$ | 21 | 0.115 | — |
| *Turning and Barrier Course* |  |  |  |  |
| Condition main effect | $F_{3,45} = 10.369$ | 3, 45 | $2.63 \times 10^{-5}$ | $\eta_p^2 = 0.41 [0.20, 1.00]$ |
| OFF vs KaDBS | $t_{45} = 4.705$ | 45 | $1.00 \times 10^{-4}$ | — |
| OFF vs cDBS | $t_{45} = 4.531$ | 45 | $1.00 \times 10^{-4}$ | — |
| OFF vs iDBS | $t_{45} = 4.406$ | 45 | $2.00 \times 10^{-4}$ | — |
| *Arrhythmicity* |  |  |  |  |
| *Stepping-in-Place Task* |  |  |  |  |
| Condition main effect | $F_{3,18.9} = 2.934$ | 3, 18.9 | 0.060 | $\eta_p^2 = 0.32 [0.00, 1.00]$ |
| *Turning and Barrier Course* |  |  |  |  |
| Condition main effect | $F_{3,41.1} = 1.997$ | 3, 41.1 | 0.129 | $\eta_p^2 = 0.13 [0.00, 1.00]$ |

Sample sizes: stepping-in-place $n = 8$ , turning and barrier course $n = 7$ (combining ellipses and figure-of-eight with task as fixed effect); arrhythmicity $n = 6$ and $n = 5$ respectively for OFF condition due to complete freezing. Post-hoc comparisons: Šidák-corrected treatment-versus-control contrasts. $\eta_p^2$ , partial eta-squared with 95% confidence intervals; KaDBS, kinematic adaptive deep brain stimulation; cDBS, continuous deep brain stimulation; iDBS, intermittent deep brain stimulation.

**Supplementary Table 2:**
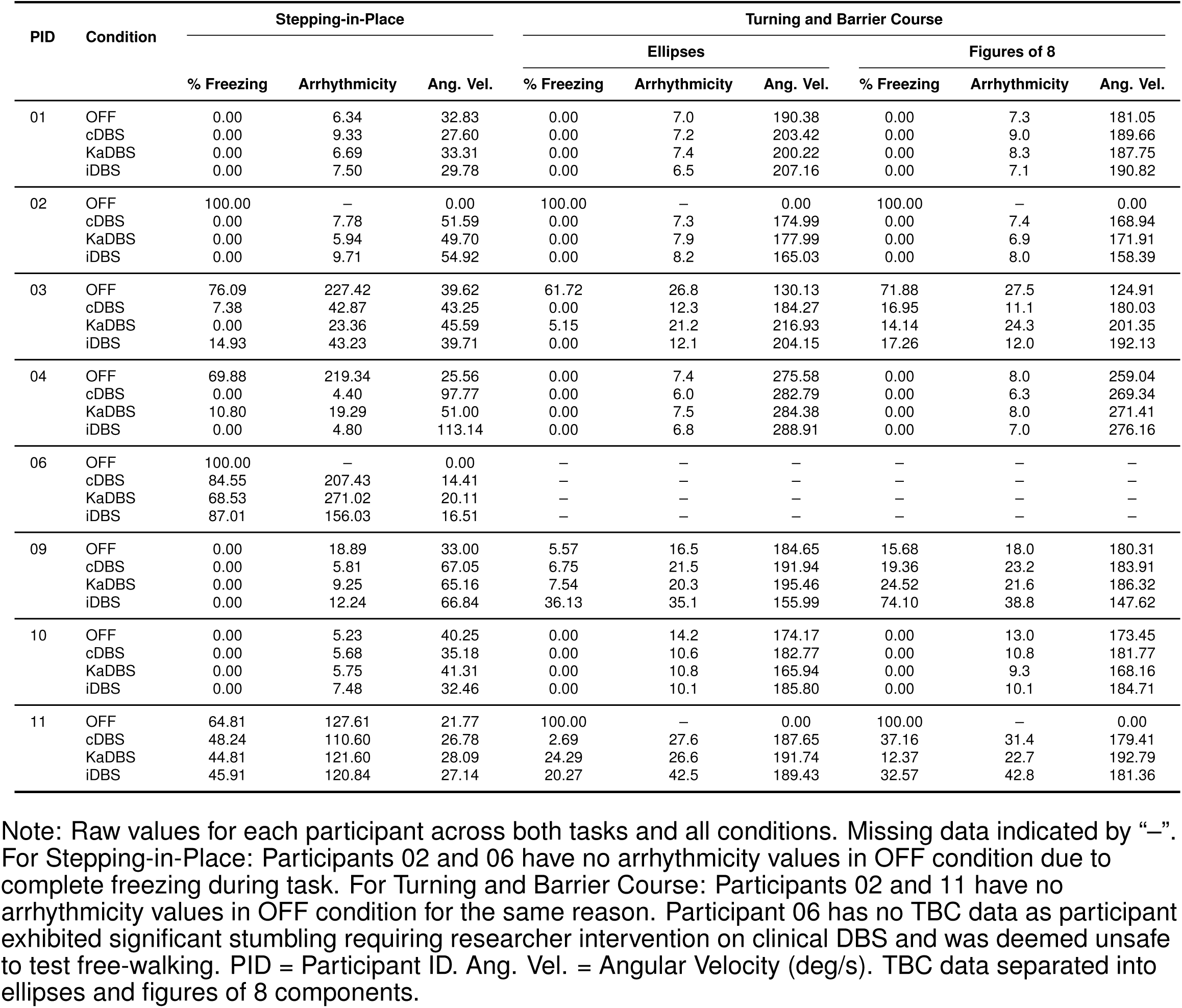
Raw Metrics by Participant and Condition.

**Fig. S2.**
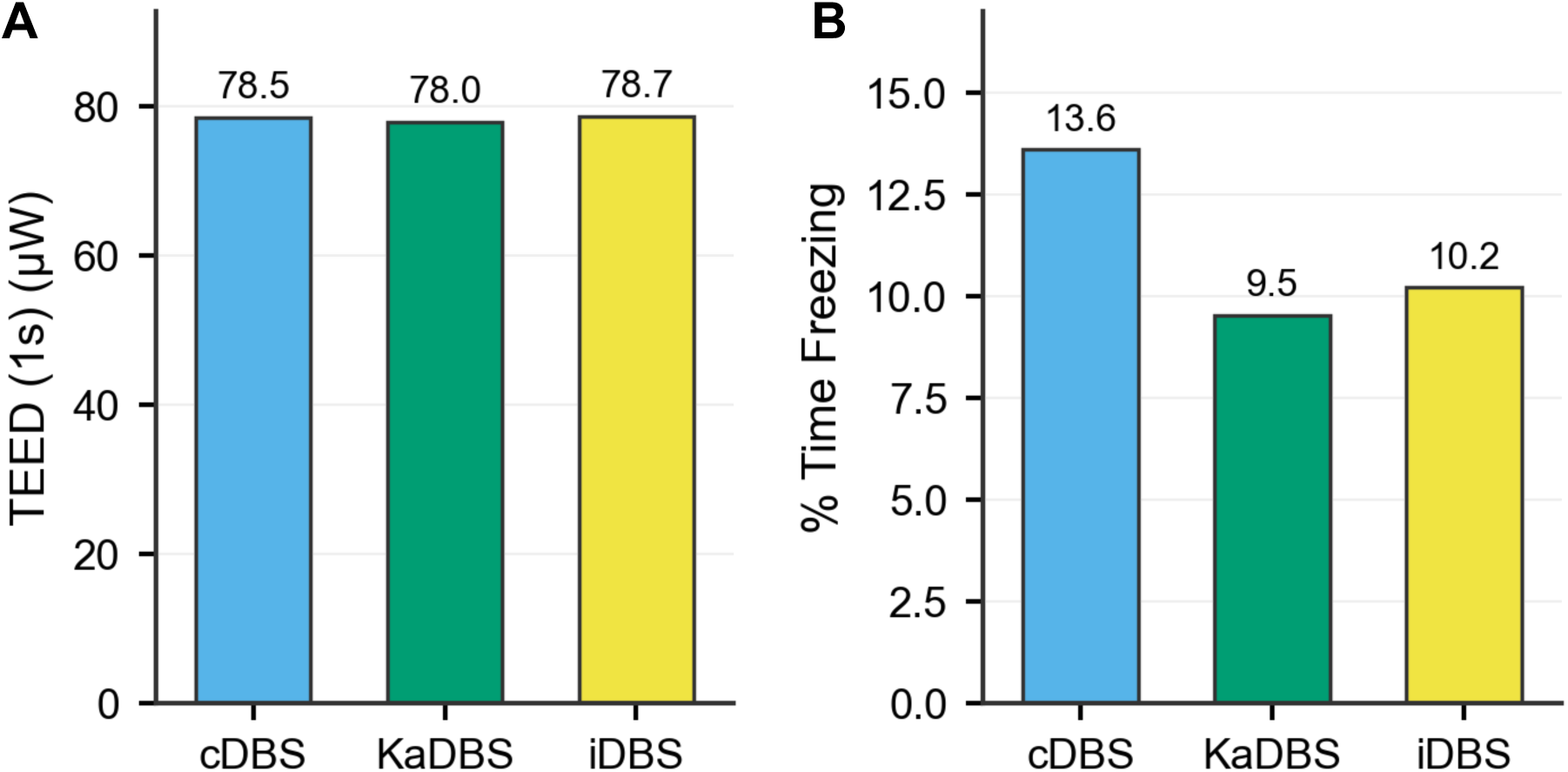
Total electrical energy delivered and freezing outcomes in exemplar participant. (A) TEED (1s) and (B) percent time freezing for Participant 03 during turning and barrier course.

**Movie S1. Arrhythmicity control algorithm modulating stimulation in real-time during stepping-in-place.** (Top) Real-time gait arrhythmicity (ARi) with participant-specific threshold ARt (red dashed line). (Middle) Controller state (Increase, Hold, Decrease). (Bottom) Bilateral STN stimulation amplitude (left, blue; right, orange) adjusted in real time during an 80 s stepping-in-place trial.

**Movie S2. P(FOG) classifier driving tri-state stimulation control during TBC.** (Top) Stepwise probability of freezing of gait, P(FOG), with dual thresholds (Pmax = 0.7, red dashed line; Pmin = 0.3, green dashed line). (Middle) Tri-state controller output (Increase, Hold, Decrease). (Bottom) Bilateral STN stimulation amplitude (left, blue; right, orange) modulated in real time during a ∼200 s turning-and-barrier course trial.

